# Improvement of Gemcitabine Treatment of Pancreatic Cancer by the Addition of All-*trans* Retinoic Acid and Identification of Vitamin A and Pentraxin 3 as Potential Response Biomarkers

**DOI:** 10.64898/2026.08.16.26359923

**Authors:** Sarah Niessen, Carola Focke, Steffen Keller, Hanna Scheffold, Silke Hempel, Johannes Lettner, Tobias Scheef, Rhena F.U. Klar, Georg Vladimirov, Kirstyn Anne Crossley, David Bittner, Max Deuter, Sandra Kissel, Sophia Chikhladze, Stefan Fichtner-Feigl, Justus Duyster, Melanie Boerries, Jakob Neubauer, Florian Scherer, Michael Lübbert, Michael Quante, Dietrich A. Ruess, Heiko Becker

## Abstract

**Background:** Therapy resistance in pancreatic ductal adenocarcinoma (PDAC) is facilitated by the desmoplastic tumor microenvironment (TME) orchestrated by cancer associated fibroblasts (CAFs). Upon activation, pancreatic stellate cells (PSCs) deplete their intracellular retinoic acid (RA)-containing lipid droplets and secrete stromal remodeling proteins like pentraxin 3 (PTX3), leading to cancer progression. Preclinical evidence indicates that all-*trans* RA (ATRA) reprograms the TME, while circulating vitamin A and PTX3 were proposed as biomarkers for ATRA response in PDAC. To support further clinical development of RA-based therapies in PDAC, we studied the effects of ATRA on CAFs and patient-derived organoids (PDO) and evaluated the clinical relevance of these biomarkers in PDAC patients.

**Methods:** We employed viability assays in human and murine organoid mono-and co-culture models to explore the efficacy of adding ATRA to gemcitabine (GEM). In parallel, we conducted a prospective observational study and assessed vitamin A and PTX3 as response biomarkers in peripheral blood collected before first treatment and at cycles 2 and 4 of treatment among patients with advanced PDAC receiving GEM with or without nab-paclitaxel (NAB-P).

**Results:** In PDO monocultures, a significant additive effect of ATRA in combination with GEM on viability was observed in 5 (41%) of 12 PDOs and this effect was numerically more frequent in organoids from patients who had clinically responded to GEM. In human and murine 3D PDO+PSC/CAF co-cultures, ATRA demonstrated an additional direct impact on the viability of stromal cells. Clinically, among 18 patients with PDAC treated with GEM+/-NAB-P, patients with no treatment response (n=10) showed an increase in PTX3 and concomitant decrease in vitamin A levels under therapy. In contrast, response was associated with stable vitamin A levels and a trend towards lower PTX3 levels during chemotherapy.

**Conclusions:** Our preclinical data support the repurposing of ATRA, an agent with favorable toxicity profile, to potentiate the efficacy of GEM in PDAC treatment. Complementing these results, our clinical data suggest vitamin A and PTX3 as promising response biomarkers in PDAC treatment, not restricted to ATRA containing regimens.

## Background

Pancreatic ductal adenocarcinoma (PDAC) is an aggressive cancer often detected at advanced stages, limiting treatment options to palliative chemotherapy regimens based on gemcitabine (GEM) or fluoropyrimidines. The prognosis of PDAC patients remains poor with a 5-year overall survival (OS) of only about 11% (Hosein et al., 2020). Therapeutic resistance contributing to the dismal prognosis associated with PDAC has been attributed to the genomic profile - with frequent mutations in the *KRAS* oncogene and in tumor-suppressor genes including *TP53, CDKN2A* and *SMAD4* - and to the immunosuppressive and desmoplastic tumor microenvironment (TME) (Beatty et al., 2021; Grünwald et al., 2021; Kung et al., 2025; Zhang et al., 2023).

The TME consists of diverse cell types, including cancer-associated fibroblasts (CAFs) and a largerly immunosuppressive immune infiltrate mainly consisting of tumor-associated macrophages (TAMs) and other myeloid cells. CAFs are a major contributor to the dense, hypoxic and desmoplastic stroma, impairing drug delivery to tumor cells. CAFs originate from multiple cell types and the complexity and heterogeneity of CAF populations can lead to diverse phenotypes, either pro-or anti-tumorigenic (Tao et al., 2025). One source of CAFs are activated pancreatic stellate cells (PSCs), which produce extracellular matrix (ECM) components in response to injury or inflammation (Helms et al., 2022; Sunami et al., 2020; Thakur et al., 2025; Zhang et al., 2022).

In healthy conditions, PSCs are quiescent and store retinoic acid (RA) in cytoplasmic droplets, which they deplete upon activation during inflammation. This activation promotes ECM production and contributes to carcinogenesis (Seidel et al., 2025; Xu et al., 2010). Retinoid signaling is essential for normal pancreas development, regulating key signaling pathways that are disrupted during pancreatic carcinogenesis (Bleul et al., 2015; Kocher et al., 2020; Sun et al., 2024). Patients with PDAC often exhibit a deficiency of fat-soluble vitamins such as vitamin A, which could sustain PSC activation fostering desmoplastic TME and therapy resistance (Froeling et al., 2011). Preclinical studies show that restoring RA in activated PSC with all-*trans* RA (ATRA) can reduce desmoplasia and cancer progression (Chronopoulos et al., 2016; Kocher et al., 2020). To date, ATRA is approved for the treatment of acute promyelocytic leukemia (APL), where it has shown safety and efficacy to treat this fulminant disease (Lo-Coco et al., 2013). In PDAC, Kocher et al. investigated ATRA as a stromal-targeting agent combined with GEM and nab-paclitaxel (NAB-P) in a phase I trial (STARPAC, NCT03307148). They demonstrated safety of the combination and potential therapeutic benefit. They also suggested vitamin A and pentraxin 3 (PTX3) blood levels as biomarkers for ATRA response (Kocher et al., 2020).

PTX3 is produced at sites of inflammation by fibroblasts, macrophages or neutrophils where it regulates inflammation, tissue remodeling and cancer progression (Doni et al., 2019). Elevated serum PTX3 has shown higher diagnostic accuracy than routine parameters (CA19-9 or CEA) for detecting PDAC, particularly in clinically ambiguous cases (Goulart et al., 2021; Zajkowska and Mroczko, 2023). A meta-analysis also demonstrated an association between elevated serum PTX3 and poorer survival in different malignancies, including PDAC (H. Jung et al., 2024). However, data from the SCALOP trial suggested that PTX3 is not prognostic in patients receiving radiochemotherapy for locally advanced PDAC (Goulart et al., 2021). To date, no study analyzed PTX3 levels - along with vitamin A levels - as response biomarkers to standard treatment in advanced PDAC.

In summary, the favorable toxicity profile of RAs, the strong preclinical evidence for TME modulation and the poor prognosis of patients with PDAC strongly support the further development of RA combination therapies in PDAC. Thus, to inform the clinical development of RA-based trials, we assessed the effects of ATRA in PDO and PDO+PSC/CAF co-cultures, and we prospectively evaluated vitamin A and PTX3 as biomarkers in PDAC patients receiving standard GEM-based therapy, without ATRA as stromal-targeting treatment.

## Methods

### Patient-Derived Tissue Collection and Cell Lines

PDAC tissue samples, either from the primary tumor or from metastasis, were collected from patients undergoing surgical exploration and resection at the Medical Center University of Freiburg between 2016 and 2022. All patient samples were obtained with informed consent. The study was approved by the local ethics committee (reference 126/17). Patient tumor specimens were immediately transported to the laboratory for further processing. Two independent human fibroblast lines (SV1794, CT1385) were isolated from patient-derived tumor tissue using the outgrowth method and served as representative, non-patient-matched CAF models, as matched fibroblast lines were not available for each individual PDO. Murine PDAC cell lines and organoids were derived from genetically engineered KPC compound mutant mice (*Kras^tm1Tyj^*: *Kras^LSL-G12D/+^*+ *Trp53^tm1Brn^:Trp53^fl/fl^* + *Ptf1a^tm1(Cre)Hnak^: Ptf1a^Cre-ex1/+^).* These mice spontaneously develop PDAC and represent a well-established model for studying tumor-stroma interactions and disease progression (Lee et al., 2016). Animals were housed at the facilities of the Center for Experimental Models and Transgenic Services (CEMT) at the Medical Center, University of Freiburg, Germany, under hygienic, pathogen-free conditions, and all animal procedures were approved by the local authority (Regierungspräsidium Freiburg, Germany; approval number G/19-137). Genotyping was performed after weaning and post mortem.

### Drug Testing in PDO Mono-Culture

Establishment and maintenance of PDO cultures was performed as previously described (Baker et al., 2016; Hafner et al., 2026; Rittmann et al., 2021), with slight adaptations. PDOs were cultured in 3D Matrigel cultures and exposed to GEM (MedChem (HY-17026); 0.5-10nM) and ATRA (MedChem (HY-14649)), either as single agents or in combination. Based on previously conducted titration experiments and literature (Koikawa et al., 2021), a fixed concentration of 10 µM ATRA was used for all subsequent experiments. Treatment was maintained for 5 days. Afterwards cell viability was assessed using the CellTiter-Glo® 3D Cell Viability Assay (Promega) following the manufacturer’s protocol. After treatment, 100 µL of CellTiter-Glo 3D reagent was added to each well. Plates were shaken at 800 rpm for 5 minutes and then incubated at room temperature (RT) for an additional 10-15 minutes to stabilize the luminescent signal. Luminescence was measured using a Tecan microplate reader. Signal intensities were background-subtracted and normalized to DMSO-treated control samples. To better visualize the additive effect of ATRA, DMSO-normalized viability was further normalized to viability under GEM monotherapy and defined as relative viability.

### Drug Testing in 3D Co-Culture

PDOs were established as described above. Murine PDAC organoids were derived from KPC mice. CAFs (or PSCs only, from healthy mouse littermates) and PDAC organoid forming units (OFU) were harvested and prepared according to standard passaging protocols. For co-cultures, fibroblasts and OFUs were mixed at a 2:1 ratio, using 2000 fibroblasts and 1000 OFUs per 5 µL dome. The cell suspension was embedded in Collagen Type I (rat tail, ibidi, #50201) and seeded into 96-well plates in 5 µL domes (in triplicate). Following 20 minutes of polymerization at 37 °C, 100 µL of standard culture medium was added per well. After 24 hours, cultures were treated as described above. For co-culture assays, 1000 to 3000 cells or OFUs were embedded in 5 µL of Collagen I per well. Signal intensities were background-subtracted and normalized to DMSO-treated control samples.

### Biomarker and response assessment in a prospective observational study

We conducted a prospective, observational cohort study. All patients included in this analysis had histologically confirmed, advanced PDAC and were planned for GEM-based chemotherapy (GEM monotherapy or GEM+NAB-P) according to physician’s recommendation. The study was approved by the ethics committee of the University of Freiburg (reference 23-1226-S1). All patients had given written informed consent for collection and use of data and specimens. In brief, patients were treated with GEM 1000 mg/m^2^ on days 1, 8, and 15 ± NAB-P 125 mg/m^2^ on days 1, 8 and 15 of a 28 days cycle, with dose reductions if deemed necessary.

Blood was collected before cycle 1 (pre-treatment) and at cycle 2 and at cycle 4 for biomarker assessment. Serum vitamin A (retinol), C-reactive protein (CRP), neutrophil to lymphocyte ratio (NLR), CEA and CA19-9 levels were measured at the hospital laboratory of the Medical Center, University of Freiburg. CRP values were excluded from further analyses if assessed during infection. For PTX3 analysis, plasma was isolated from blood samples, frozen and stored at the FREEZE Biobank of the Medical Center, University of Freiburg. Circulating tumor (ct)DNA was measured in the plasma as described below.

Clinical data were collected pre-treatment and at each timepoint. Treatment response was assessed after 2-3 cycles of chemotherapy according to physician’s choice using clinical examination, imaging (computed tomography (CT) scan or magnetic resonance imaging (MRI)), and tumor marker measurement in the serum (CA19-9 and CEA).

### PTX3 ELISA

PTX3 levels were quantified with a sandwich enzyme-linked immunosorbent assay (ELISA) in a blinded manner, using in-house validated method based on a monoclonal antibody MNB4 (Enzo Life Sciences ALX-804-464-C100). Plasma PTX3 concentrations were measured using the sandwich ELISA as follows: 96 well-ELISA plates were coated overnight at 4°C with MNB4 anti-human PTX3 antibody (2 µg/ml) diluted in coating buffer (Themofisher), washed, blocked (Thermofisher, 2 h, at RT), washed, and incubated with either 100 μL of undiluted plasma or 100 μL recombinant human PTX3 standards (0.15–20 ng/mL), all in triplicates for 2 h at 37 °C. After two washes, 50 ng/mL of biotinylated PTX3 antibody (Enzo Life Sciences, cat. ALX-210-365B) was added in each well for 1 h at RT. The plates were washed and color realized by 100 μL/well streptavidinhorseradish peroxidase (Thermo Scientific) diluted 1:4000 for 1 h at RT. After further washes 100 μL of chromogen substrate (ThermoFisher cat. 34028B) was added and after 15 min, 50µL of stop solution (1M H2SO4) was added to each well to stop the reaction. Plates were immediately read at 450 nm in a plate-reader. Polynomial regression graphs were constructed for standard curves (**Supplementary figure (fig.) 7**). Plasma samples of each patient time-point were thawed only once and assayed directly, maintaining a chain of custody, in triplicate. Patient variables were unblinded after submission of readouts.

### Circulating tumor DNA measurements

In patients with known mutational status, which was determined by routine molecular pathology diagnostics of tumor tissue, we measured ctDNA in plasma before and during GEM-based chemotherapy at the Liquid Biopsy Laboratory of the Medical Center, University of Freiburg, as described previously (Hussung et al., 2021). Briefly, cell-free DNA (cfDNA) was isolated from plasma, and the sequence variant was analyzed with a digital Droplet-PCR (ddPCR) assay. The allele frequency was calculated as followed: number of mutated copies per ml plasma / (number of mutated copies / ml plasma + number of wild type copies / ml plasma) * 100%.

### Body composition measurements

Body composition was assessed using abdominal CT scans routinely performed before starting the GEM-based treatment. CT images were processed using NORA software for body composition analysis as described before (M. Jung et al., 2024). Areas of skeletal muscle (SM), visceral adipose tissue (VAT), subcutaneous adipose tissue (SAT) and intramuscular adipose tissue (IMAT) were quantified on CT images at the level of the 3rd lumbar vertebral body. The software automatically differentiated these regions based on pre-set Hounsfield Unit (HU) thresholds, with manual adjustments made when necessary to ensure accurate delineation. Parallel to body composition analysis we assessed for each patient and timepoint weight and body mass index (BMI).

### Statistical analyses

Sample size considerations for PDO models and biomarker assessment were based on feasibility considerations. Statistical analysis and graphical data representation were performed using GrahPad Prism version 10 (GraphPad Software, LLC, Boston, Massachusetts, USA). Data are given as mean ± standard deviation (SD). Statistical tests are described in figure legends. Dose-response curves of PDO viabilities were analyzed by nonlinear regression using a four-parameter logistic model. To statistically compare assay results of human and murine PDO mono-and co-cultures we used two-way ANOVA.

For the prospective observational cohort, as the sample size was small, non-parametric tests were applied. Comparisons between two independent groups (responders vs. non-responders) at each time point were performed using the Mann-Whitney test. To control the family-wise error rate arising from multiple testing, p-values were adjusted using the Holm-Šídák method. Longitudinal changes within each group were assessed using pairwise comparisons between two time points with the Wilcoxon signed-rank test for paired data. Simple linear regression was performed to assess the relationship between vitamin A and time for each group. Slopes were compared between groups using Prism‘s built-in slope comparison test (GraphPad Software, LLC, Boston, Massachusetts, USA). The body composition analysis was performed using Welch’s t-test. All statistical tests were two-sided, and a p-value < 0.05 was considered statistically significant.

## Results

### Gemcitabine in combination with ATRA in PDAC patient-derived organoid models

ATRA-based therapy is highly effective in APL (Lo-Coco et al., 2013). Beyond inducing differentiation, it can reverse aberrant epigenetic programs and modulate cancer-associated chromatin (Arteaga et al., 2015; Meier et al., 2022), thereby potentially influencing drug sensitivity in cancer models (Tang and Gudas, 2011). While ATRA has mainly been studied for stromal modulation in PDAC (Carapuça et al., 2016; Chronopoulos et al., 2016), we hypothesized that ATRA also exerts tumor cell-intrinsic effects that alter chemosensitivity.

We first assessed the combined effect of ATRA and GEM on the viability of 12 PDO lines established from tumors of 11 PDAC patients. Organoids from either primary tumor (PT), peritoneal (PC) or liver metastasis (LM), were treated with increasing concentrations of GEM, either alone or in combination with ATRA. In viability assays, co-treatment with GEM and ATRA significantly reduced cell viability compared to GEM alone in selected GEM concentrations and PDO lines (**Fig. 1**). This suggests that ATRA may potentiate the effects of GEM in PDAC PDOs and indicates that tumor cell-intrinsic effects may also contribute to the therapeutic potential of ATRA in PDAC. A significant additive effect of ATRA in combination with GEM was observed in 5 (41%) of 12 PDOs.

**Figure 1.**
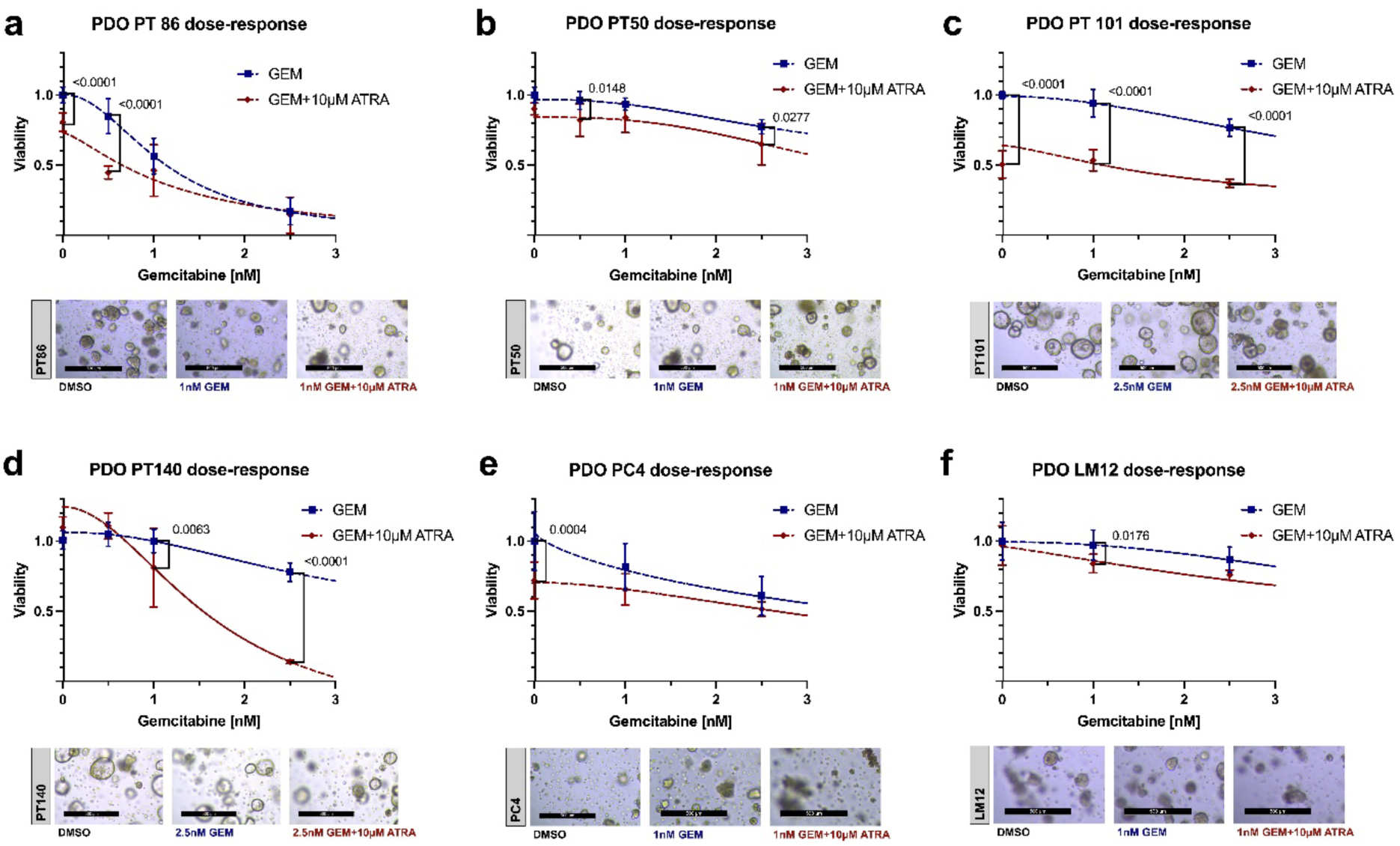
Effect of ATRA + GEM on PDO Viability. Dose-response curves showing combined effect of GEM and ATRA on cell viability of 6 PDAC PDOs following treatment with GEM alone or in combination with 10 µM ATRA. Individual panels represent distinct organoid models: **a)** PDO PT86, **b)** PDO PT50, **c)** PDO PT101, **d)** PDO PT140, **e)** PDO PC4 and **f)** PDO LM12. Organoids were treated with increasing concentrations of GEM as indicated. Viability was normalized to DMSO-treated controls. Data points represent mean ± SD. Dashed lines indicate nonlinear regression fits generated using a dose-response inhibition model. Statistical analysis was performed using two-way ANOVA comparing GEM versus GEM+ATRA treatment at individual GEM concentrations; significant differences are indicated in the graphs. Representative brightfield images of organoids treated with DMSO, GEM alone or GEM + ATRA at the indicated concentrations are shown below each corresponding dose–response plot. Scale bars, 500 µm.

The effect was more frequent in organoids from patients with response to GEM treatment. Response data were available for 10 of the 11 patients from whom PDOs were established. Among four patients with response to GEM treatment, PDOs from three patients (75%) exhibited an additive *in vitro* response to GEM+ATRA. In contrast, an additive response was observed in PDOs from two of six patients (33%) who did not respond to GEM (**Supplementary Table 1**). For one patient, we analyzed two spatially distinct PDOs (LM5&PC4) and observed different dose response curves, albeit there was no significant difference in the viability responses between GEM+ATRA and GEM alone in both PDOs (**Supplementary Table 1 and Fig. 1)**, underscoring the impact of intertumoral heterogeneity on therapy response.

### Gemcitabine in combination with ATRA in human and murine 3D co-culture models

While ATRA is known to reprogram CAFs/PSCs and to modulate tumor-stroma interactions (Carapuça et al., 2016; Sun et al., 2024), we aimed to examine its potential indirect effects on tumor cell viability. Thus, to account for the role of the TME, particularly CAFs, in ATRA therapy response, we evaluated the therapeutic effects in four independent 3D co-culture models (n=2 human and n=2 murine) comprising CAFs or PSCs, compared with the respective PDO and CAF/PSC monocultures (**Fig. 2**).

**Figure 2.**
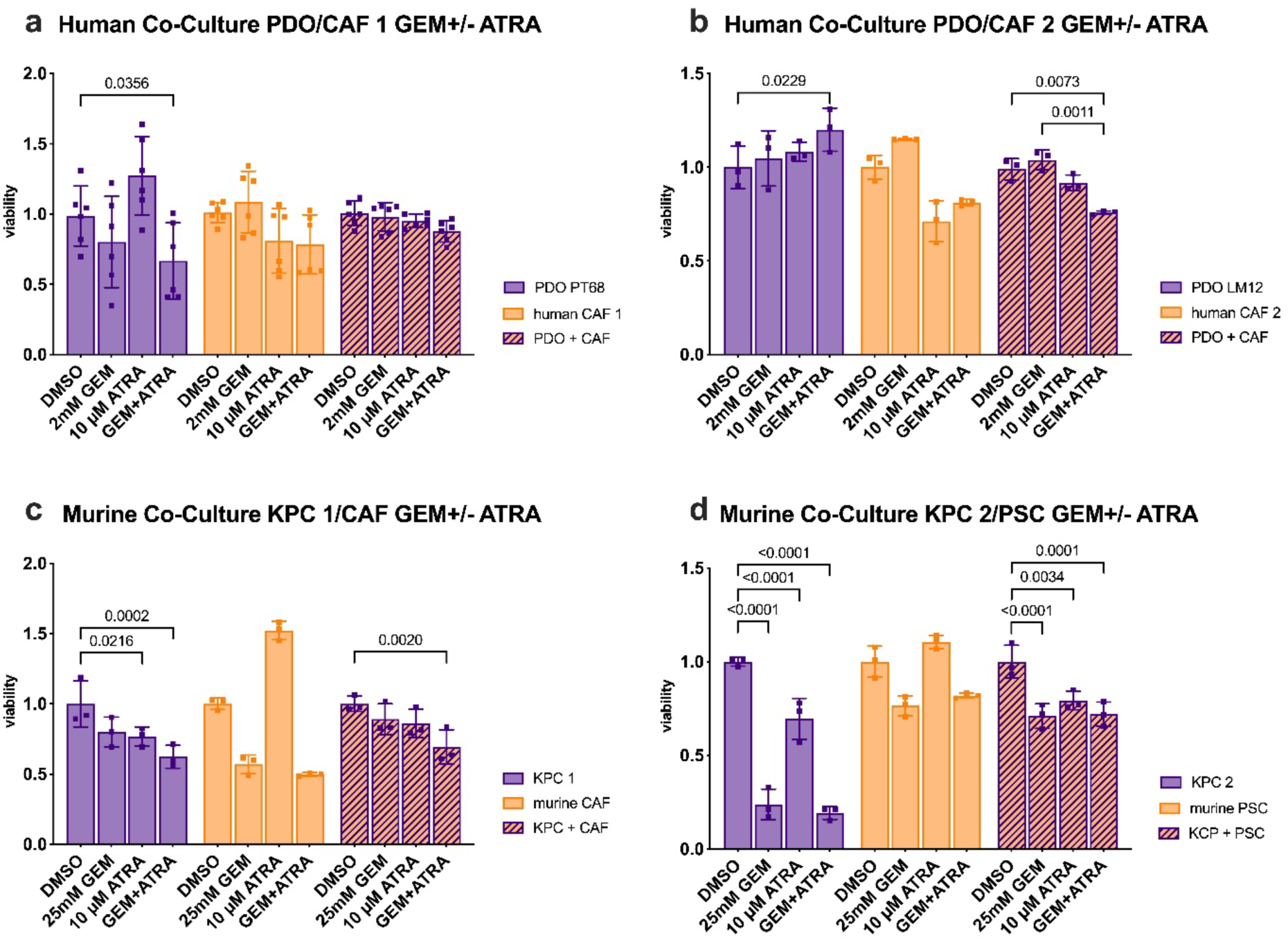
ATRA + GEM in a co-culture model of human and murine PDAC organoids with CAFs. Cell viability in **a)+b)** human and **c)+d)** murine 3D mono-and co-cultures of PDAC cells and cancer-associated fibroblasts (CAFs) or pancreatic stellate cells (PSCs) following treatment with ATRA (10 µM) and GEM. Purple represents 3D PDAC monoculture, orange 3D CAF/PSC monoculture, and striped 3D PDAC/CAF co-culture. GEM concentrations are indicated in the graph titles. Each data point represents the mean of a technical triplicate; error bars indicate the SD of these triplicates. In panel b) co-cultures show a significant decrease in viability upon combined treatment with ATRA and GEM compared to GEM alone (two-way ANOVA). Only significant p-values are shown.

In CAF co-culture models the effect of co-treatment with GEM and ATRA on viability differed depending on the stromal cell context. A significant reduction in cell viability compared to GEM alone was only observed in the LM12/SV human co-culture model (**Fig. 2b**). Notably, we also observed effects of the combination treatment compared to DMSO on viability in human fibroblast 3D monocultures (**Fig. 2a and 2b**), indicating a direct impact of ATRA on stromal cells, which we interpret as changes in cellular activity or proliferation, rather than cell death. These findings support the hypothesis that ATRA may enhance the efficacy of chemotherapy by modulating CAFs within the TME.

Together with the PDO monoculture data, these findings suggest that ATRA could enhance GEM efficacy via stromal-mediated effects. **However, the heterogeneous responses observed across the co-culture models preclude firm conclusions regarding the relative contribution of stromal-mediated versus tumor cell-intrinsic mechanisms.**

### Patient and disease characteristics of the prospective, observational cohort

To inform future clinical trials of GEM+RA combination therapy beyond preclinical evidence, we aimed to evaluate potential biomarkers of response to standard GEM therapy, that may also be relevant in the context of GEM+RA combination therapy.

For this purpose, we prospectively assessed a panel of biomarkers in the peripheral blood from advanced PDAC patients prior to cycle 1 and at cycles 2 and 4 of a GEM-based chemotherapy, without ATRA. We enrolled 18 patients; 89% had metastatic disease, and 72% received GEM+NAB-P, the remaining GEM monotherapy. Fourteen (78%) patients harbored a *KRAS* mutation. The *TP53* mutation status was available for 17 patients, 76% of whom had a mutation. *CDKN2A* and *SMAD4* were each mutated in 36% of 14 patients analyzed (**Table 1**, **Fig. 3**).

**Figure 3.**
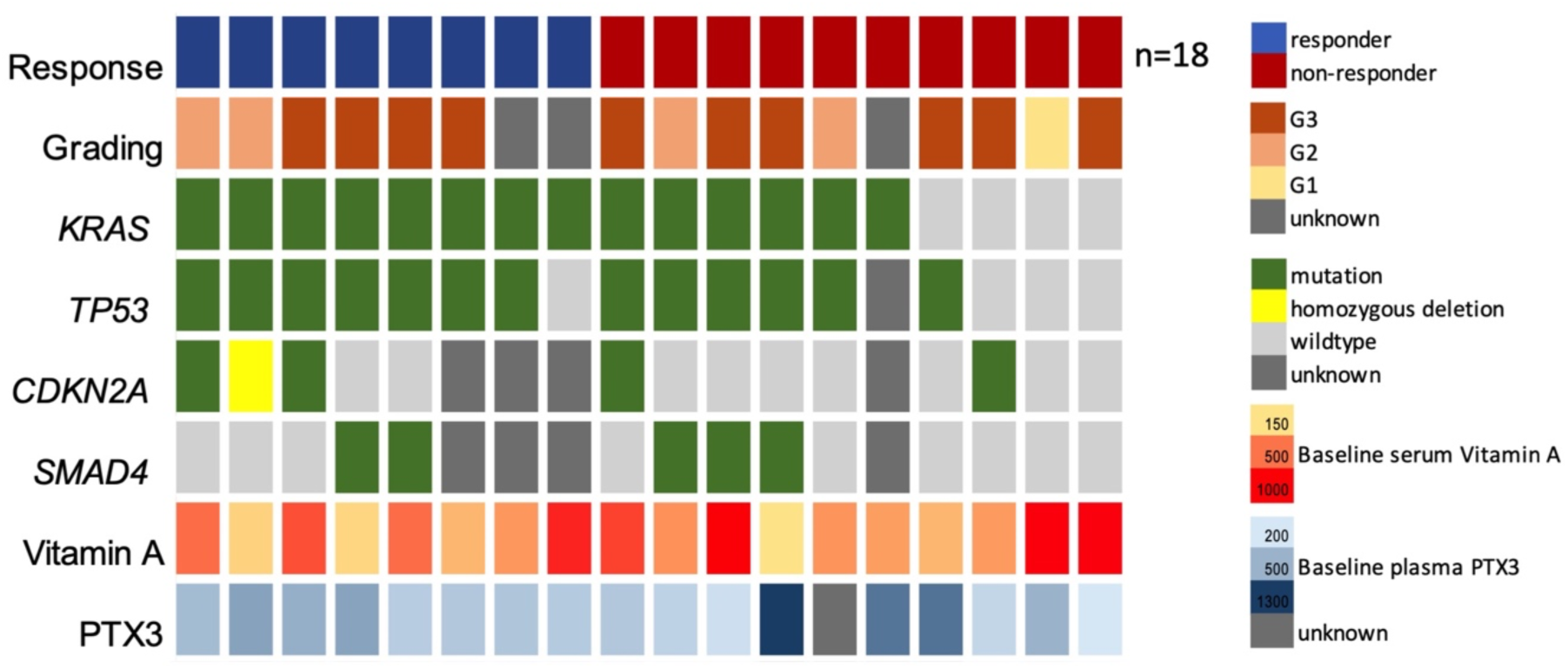
Case-level profile of 18 PDAC patients. Each column represents one case. Each row represents one clinical characteristic or mutation or biomarker. Baseline serum vitamin A levels are shown as color gradient, with light yellow color representing lower levels and red color high levels. Similarly baseline plasma PTX3 levels are shown as blue color gradient, with light color representing lower and dark blue color higher levels. Darker Grey color represents unknown values. Comparisons of vitamin A and PTX3 levels between responders vs. non-responders at baseline were performed using the Mann-Whitney test.

**Table 1.** Patient and disease characteristics for prospective response biomarker assessment.

| Characteristics | No. (%) |
| --- | --- |
| **All patients** | 18 (100%) |
| **Sex** |  |
| m | 11 (61%) |
| f | 7 (39%) |
| **Mean age at first diagnosis (years)** | 67 (range 50-79) |
| **Stage at diagnosis** |  |
| Localized | 10 (55%) |
| Metastasized | 8 (45%) |
| **Stage at study inclusion** |  |
| Localized | 2 (11%) |
| Metastasized | 16 (89%) |
| **Histology** |  |
| Ductal adenocarcinoma | 16 (89%) |
| IPMN with associated invasive carcinoma | 2 (11%) |
| **Differentiation** |  |
| Well (G1) | 1 (5%) |
| Moderate (G2) | 4 (23%) |
| Poor (G3) | 10 (55%) |
| Unknown | 3 (17%) |
| **Therapy during biomarker assessment** |  |
| Gemcitabine monotherapy | 5 (28%) |
| Gemcitabine + nab-paclitaxel | 13 (72%) |
| **Previous lines of therapy prior to Gemcitabine-based treatment** |  |
| No | 8 (45%) |
| Yes | 10 (55%) |
| **Treatment intention during biomarker assessment** |  |
| Palliative | 17 (95%) |
| Neoadjuvant | 1 (5%) |
| **Molecular markers (patients with mutations/patients analyzed)** |  |
| KRAS mutation | 14/18 (78%) |
| TP53 mutation | 13/17 (76%) |
| CDKN2A mutation | 5/14 (36%) |
| SMAD4 mutation | 5/14 (36%) |

To clearly distinguish patients with treatment response, responders had to fulfill the following criteria: stable disease or better in CT/MRI and >30% reduction in tumor marker levels from baseline and a decrease in plasma ctDNA after one cycle of chemotherapy, if measurable. These criteria considered those used in the STARPAC trial to define biochemical responders (Kocher et al., 2020).

A treatment response was observed in 8 patients (44%, **Fig. 3**). All other patients were classified as non-responders, i.e. patients with progressive disease in CT/MRI, or no sufficient reduction of tumor marker level, or increase of ctDNA in plasma, or death. Baseline PTX3 and vitamin A values did not significantly differ between responders and non-responders (**Fig. 3**).

### PTX3 and vitamin A blood levels during gemcitabine-based therapy

PTX3 is known as a general marker for PDAC detection and possibly prognosis (Goulart et al., 2021; H. Jung et al., 2024; Sato et al., 2022). Vitamin A (retinol) can be oxidized to RA, the transcriptionally active form (Blaner, 2019; Carazo et al., 2021). Although retinoid signaling is essential for pancreas biology, vitamin A has not been studied as a general marker for PDAC detection or prognosis. In the STARPAC trial, PTX3 and vitamin A were proposed as potential response markers to ATRA treatment in PDAC (Kocher et al., 2020). However, their role as biomarkers for response to standard treatment, independent of ATRA, remains unclear. Given that PTX3 is a general marker for PDAC (Goulart et al., 2021; Watt et al., 2014), not specific for ATRA treatment, and that vitamin A is biologically distinct from ATRA, we hypothesize that both markers could also serve as response biomarker to standard treatment.

In our prospective observational cohort of advanced PDAC patients undergoing chemotherapy, responders showed a non-significant downward trend in PTX3 levels and stable vitamin A levels over time (**Fig. 4a, b, d-f, Supplementary Fig. 2a**). In contrast, PTX3 levels increased among non-responders, with the difference between non-responders and responders becoming significant at cycle 4 of chemotherapy (adjusted *P*=0.0283, **Fig. 4a**). Moreover, non-responders exhibited significantly lower vitamin A levels after one cycle of chemotherapy compared with baseline (**Fig. 4c**), although vitamin A levels were not significantly different between responders and non-responders throughout the treatment course (**Supplementary Fig. 2b**). Simple linear regression of vitamin A over time showed no significant difference in slopes between groups; however, non-responders displayed a downward trend, whereas responders showed a slight increase (**Fig. 4e**).

**Figure 4.**
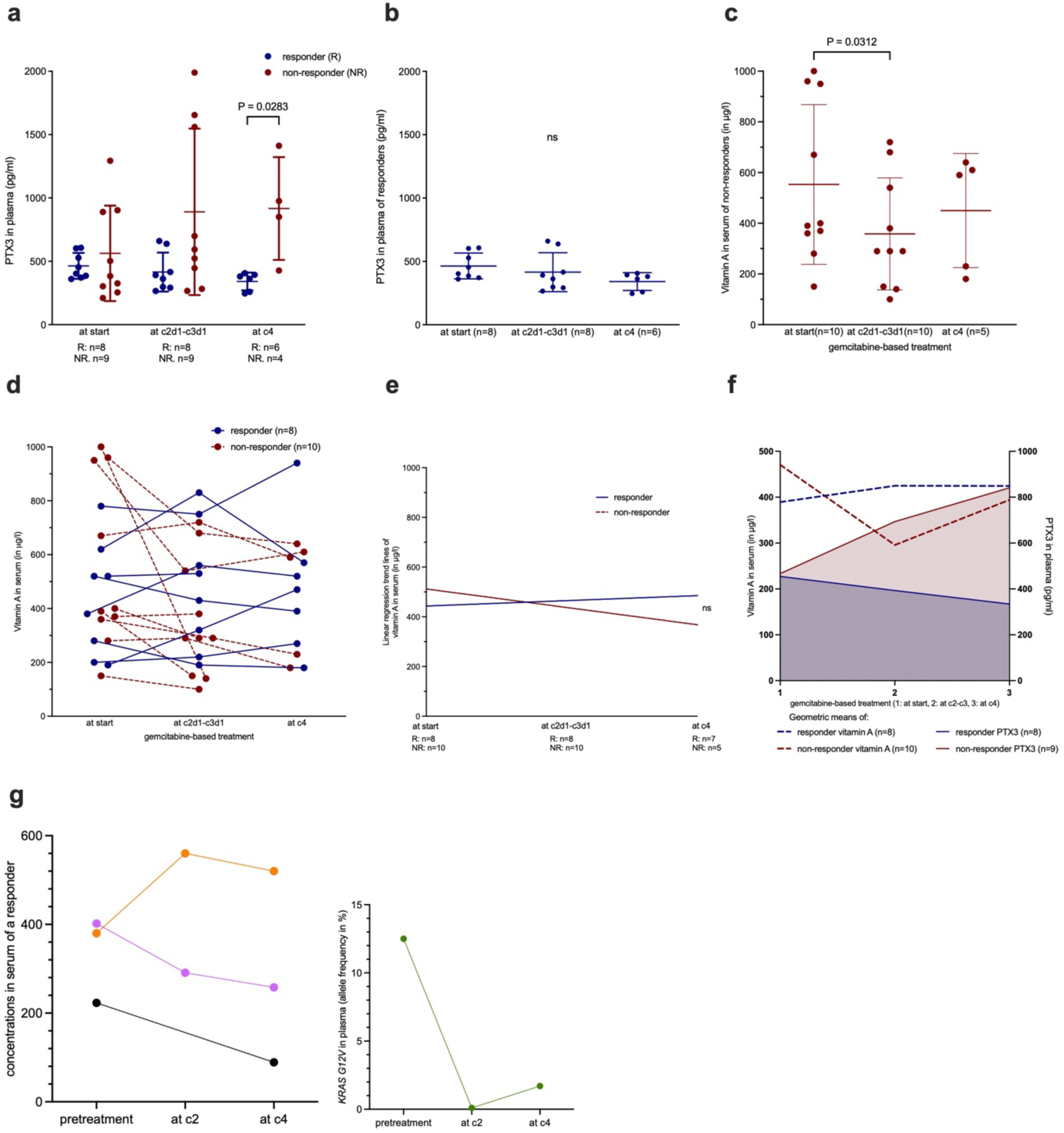
PTX3 and vitamin A in plasma of PDAC patients undergoing GEM-based treatment. **a)** Plasma PTX3 levels during the first four cycles GEM-based therapy in patients with treatment response (blue dots, n=8 at start) versus non-response (red dots, n=9 at start), measured by ELISA. Each data point represents the mean value of three readings. Data are expressed as mean and SD. Statistics were performed with multiple Mann-Whitney tests and adjusted using the Holm-Šídák method. **b)** Plasma PTX3 levels in responders to GEM-based treatment. Each data point represents the mean value of three readings. Data are expressed as mean and SD. Statistics were performed with Wilcoxon test. **c)** Serum vitamin A levels during the first four cycles of GEM-based therapy in non-responders. Each data point represents single vitamin A measurement. Data are shown as mean and SD. Statistics were performed with Wilcoxon test. **d)** Individual serum vitamin A levels during the first four cycles of GEM-based treatment. Blue dots represent responders (n=8 at start), red dots non-responders (n=10 at start). For each patient, values from different timepoints are connected with lines. **e)** Simple linear regression trend lines comparing serum vitamin A levels of responders (blue line, R squared 0.006, slope best-fit value 20.96) to non-responders (red dashed line, R squared 0.042, slope best-fit value-72) during the first four cycles chemotherapy. The slopes, elevations or intercepts are not significantly different. **f)** Geometric means of serum vitamin A levels (left axis, dashed lines) and plasma PTX3 levels (right axis, full lines and filled area under the lines) during therapy in responders (blue) and in non-responders (red). X-axis represents the time: 1: pre-treatment; 2: at c2; 3: at c4. Geometric mean was calculated from log-transformed data using back-transformation in GraphPad Prism. **g)** Representative case showing biomarker dynamic during the first four cycles of GEM monotherapy in a patient with response.

PTX3 and vitamin A have both been linked with inflammation (Carazo et al., 2021; Liu et al., 2025). Consistently, non-responders tended to have higher CRP serum levels and NLRs (neutrophil to lymphocyte ratio) than responders, but the differences were not significant at any time point (**Supplementary Fig. 3a, b**).

These results, albeit in a small cohort, support PTX3 and vitamin A as potential response biomarkers in PDAC patients treated with GEM-based treatment, with increasing plasma PTX3 levels and a trend toward decreasing serum vitamin A levels potentially indicating lack of treatment response (**Fig. 4f**).

Since dynamics of vitamin A and PTX3 levels may rather reflect changes in the TME, we correlated them with ctDNA levels as a surrogate marker for tumor cell mass itself. We studied ctDNA in 12 patients with a *KRAS* mutation and one patient with a *BRAF V600E* mutation (**Supplementary Table 2**). Notably, all responders with detectable ctDNA at baseline showed a decrease in ctDNA values after one cycle chemotherapy (**Supplementary Fig. 4a and b**).

**Fig. 4g** illustrates the informative course of the various biomarkers assessed in a patient with treatment response: decrease of ctDNA, decrease of CA19-9, decrease in plasma PTX3 levels and an increase in serum vitamin A levels.

### Body mass index and body composition during gemcitabine-based therapy

Vitamin A must be obtained by diet (Carazo et al., 2021). Therefore, we assessed whether vitamin A changes during chemotherapy were related to weight loss, indicating potential malnutrition. However, all patients had stable BMI and body weight during chemotherapy (**Supplementary Fig. 5a and Supplementary Fig. 6e**). Notably, responders generally had normal BMI levels at baseline, while obesity (BMI>30 kg/m^2^) or underweight (BMI<18.5 kg/m^2^) were more common among non-responders (**Supplementary Fig. 5a**).

To further evaluate whether vitamin A changes were associated with malnutrition, we determined the body composition from baseline CT scans (**Supplementary Fig. 5 b, c**). Quantification revealed no significant baseline difference for intramuscular adipose tissue (IMAT), subcutaneous adipose tissue (SAT) and visceral adipose tissue (VAT) between responders and non-responders (**Supplementary Fig. 5 d-f**). However, responders had narrower distributions around more physiological IMAT, SAT and VAT values, whereas non-responders showed greater variability and extreme values in IMAT, SAT, and VAT, consistent with the BMI findings (**Supplementary Fig. 5 d-f**). Again in line with the BMI data, SM (skeletal muscle) area, as well as the other body composition parameters remained stable over the course of treatment (**Supplementary Fig. 6a-d**), indicating that the observed changes in vitamin A levels are unlikely to be driven by malnutrition.

## Discussion

Patients with advanced-stage PDAC have a dismal prognosis. Novel therapeutic approaches are urgently needed. Such approaches must account for the frequently present frailty of PDAC patients, driven by older age, comorbidities or the cancer itself. RAs have a favorable toxicity profile, and their potential impact on PDAC has been demonstrated in preclinical models. To further inform the design of clinical trials evaluating RA-based therapies in PDAC, we studied the effects of ATRA in PDOs and assessed potential response biomarkers in a prospective cohort study receiving standard chemotherapy.

The additive effects of ATRA and GEM that we observed in patient-derived PDAC organoids and 3D co-culture models provide preliminary *in vitro* evidence supporting the combination of ATRA with conventional chemotherapies in humans. Our data suggest that ATRA may enhance the GEM efficacy across a range of concentrations. Importantly, additive effects were also observed in tumor-only PDO mono-cultures, indicating that ATRA activity is not solely mediated through stromal components. Beyond its effects on CAFs and PSCs, recent work in breast cancer demonstrated that ATRA modulates differentiation and proliferation via gene expression and epigenetic reprogramming, supporting a broader relevance of these mechanisms also in solid tumors (Yahyapour et al., 2025). To better capture the complexity of the TME, we evaluated ATRA and GEM in both human and murine 3D co-culture models with CAFs/PSC, which confirmed additive effects; the viability-based readouts used do not allow discrimination as to whether the observed effects are driven by treatment impacts on tumor cells, stromal compartments, or both.

Notably, the additive effect appeared most pronounced in organoids derived from patients with GEM-responsive PDAC, further supporting the investigation of ATRA as an adjunct to GEM particularly in GEM-sensitive PDAC. However, the lack of additive effects in certain organoid lines underscores the intertumoral heterogeneity in treatment response; such variability in chemotherapy efficacy is a major contributor to poor survival outcomes (Neoptolemos et al., 2018). This highlights the need for predictive biomarkers and para-clinical functional assays to identify patients most likely to benefit from this combination. In this context, PDOs and 3D co-culture systems have been proposed as valuable platforms for individualized therapy selection in PDAC (Schuth et al., 2022; Tiriac et al., 2018).

Taken together, our findings provide a strong rationale for the further clinical evaluation of ATRA in combination with GEM-based regimens in patients with advanced PDAC. The phase I trial STARPAC has demonstrated the safety of ATRA in combination with GEM + NAB-P in patients with unresectable PDAC. In this trial, PTX3 and vitamin A were suggested as biomarkers for response to ATRA treatment (Kocher et al., 2020). However, given the biological background of vitamin A and PTX3 in the carcinogenesis of the pancreas and since vitamin A (retinol) and ATRA are distinct molecules, with RA being formed through a two-step oxidation of retinol (Blaner, 2019), we hypothesized that Vitamin A and PTX3 are general biomarkers for GEM response.

Indeed, our clinical data indicate that vitamin A has value as a response biomarker for GEM-based treatment - independently of ATRA treatment. Stable serum vitamin A levels during chemotherapy indicate response, whereas non-responders may be identified by a downregulation of vitamin A after one cycle chemotherapy. Consistent with this, non-responders to ATRA+GEM+NAB-P in the STARPAC trial also showed a decrease in serum vitamin A levels during treatment (Kocher et al., 2020). Our cohort further expands on this by evaluating the potential association of vitamin A levels with weight loss or changes in body composition as marker for malnutrition - frequently present in PDAC patients - for which no associations were found. In addition, we observed that PTX3 showed a non-significant downward trend in the plasma of responders to GEM-based treatment. In contrast, non-responders exhibited elevated plasma PTX3 levels during chemotherapy, indicating increased CAF/PSC activity. The inverse trajectories of PTX3 and vitamin A levels in our cohort, especially in case of non-response, is in line with previous observations, where PDAC patients generally showed higher PTX3 and lower vitamin A serum levels than healthy subjects (Goulart et al., 2021). Thus, we propose that serum vitamin A and PTX3 are biomarkers reflecting TME remodeling during chemotherapy, also in the absence of ATRA.

Two additional findings of our study worth mentioning are the rapid decrease in ctDNA in responders within the first chemotherapy cycle, and the observation that responders exhibited narrower distributions around more physiological IMAT, SAT, and VAT values. Both observations warrant further investigation in future cohorts.

In general, our study is limited by the relatively small number of PDO PDAC models tested for treatment effects and the limited number of patients analyzed for vitamin A and PTX3. In addition, the absence of patients treated with GEM+ATRA as a comparator in the biomarker analyses, due to lack of regulatory approval, further constrains interpretation of our findings. Thus, the validity of vitamin A and PTX3 as response biomarkers in advanced PDAC will need to be established in larger prospective clinical trials, both with and without ATRA as a stromal-targeting therapy.

## Conclusion

Our preclinical data support the repurposing of ATRA, an agent with a favorable toxicity profile, to enhance the efficacy of GEM in the treatment of PDAC. Complementing these findings, our clinical data identify vitamin A and PTX3 as promising response biomarkers in PDAC therapy, not limited to ATRA-containing regimens.

## List of abbreviations

APL: acute promyelocytic leukemia
ATRA: all-trans retinoic acid
BMI: body mass index
CAFs: cancer-associated fibroblasts
CDA: cytidine deaminase
CEMT: Center for Experimental Models and Transgenic Services
cfDNA: cell-free DNA
CRP: C-reactive protein
CT: computed tomography
ctDNA: circulating tumor DNA
DCK: deoxycytidine kinase
ddPCR: digital droplet polymerase chain reaction
DNMT1: DNA methyltransferase 1
ECM: extracellular matrix
ELISA: enzyme-linked immunosorbent assay Fig. Figure
FREEZE: FREiburg und MEdizinische Fakultät ZEntrum für Biobanking (Center for Biobanking, Medical Center, University of Freiburg, Faculty of Medicine, University of Freiburg, Germany).
GEM: gemcitabine
HMA: hypomethylating agents
HU: Hounsfield units
IMAT: intramuscular adipose tissue
LM: liver metastasis
MRI: magnetic resonance imaging
NAB-P: nab-paclitaxel
NGS: next generation sequencing
NLR: neutrophil to lymphocyte ratio
OFU: organoid forming units
OS: overall survival
PC: peritoneal carcinosis
PDAC: pancreatic ductal adenocarcinoma
PDOs: patient-derived organoids
PSCs: pancreatic stellate cells
PT: primary tumor
PTX3: pentraxin 3
RA: retinoic acid
RT: room temperature
SAT: subcutaneous adipose tissue
SD: standard deviation
SM: skeletal muscle
TAMs: tumor-associated macrophages
TME: tumor microenvironment
VAT: visceral adipose tissue

## Declarations

### Ethics approval and consent to participate

The study was approved by the Ethics Committee of the University of Freiburg (ethic vote number 23-1226-S1 and 126/17). All patients had given written informed consent for collection and use of data and specimens. All procedures were in accordance with the ethical standards of the responsible committee and with the Helsinki Declaration. Animal experiments were conducted at the facilities of the Center for Experimental Models and Transgenic Services (CEMT) at the Medical Center, University of Freiburg, Germany. The animals were maintained under pathogen-free conditions, and all animal procedures were approved by the local authority (Regierungspräsidium Freiburg, Germany; approval number G/19-137).

### Consent for publication

All authors approved this manuscript for publication in the Journal of Experimental & Clinical Cancer Research.

### Availability of data and materials

Due to ethical and legal considerations, patient data from this study cannot be shared publicly. Qualified researchers may request access to anonymized data by contacting the correspending authors and providing a detailed data access proposal, subject to approval by the relevant ethics committee.

### Authors’ contributions

S.N., C.F., M.Q., D.A.R., H.B. undertook conception and design of the study; S.J.K., H.S., S.H. G.V. and C.F. performed PDO experiments; S.N., C.F., H.B., D.A.R. were responsible for treatment of patients and specimen acquisition; S.N. performed biomarker analysis; T.S. and J.N. performed body composition analysis; F.S. performed ctDNA analysis. S.N., C.F., D.A.R., H.B. wrote the manuscript. M.B., S.K., H.S., J.L., T.S., K.A.C., M.D., S.C., M.L., M.Q., S.N., C.F., D.A.R. and H.B. reviewed and edited the manuscript. All authors accepted the final version of the manuscript.

## Data Availability

All data produced in the present study are available upon reasonable request to the authors.

## Acknowledgements

The authors would like to thank all reviewers for their valuable comments. We also would like to thank the Center for Biobanking, Medical Center, University of Freiburg, Germany (FREEZE Biobank), the Liquid Biopsy Laboratory of the Medical Center, University of Freiburg, Germany and the Molecular Tumor Board of the Medical Center, University of Freiburg, Germany.

## Supplemental data

**Supplementary Table 1.**
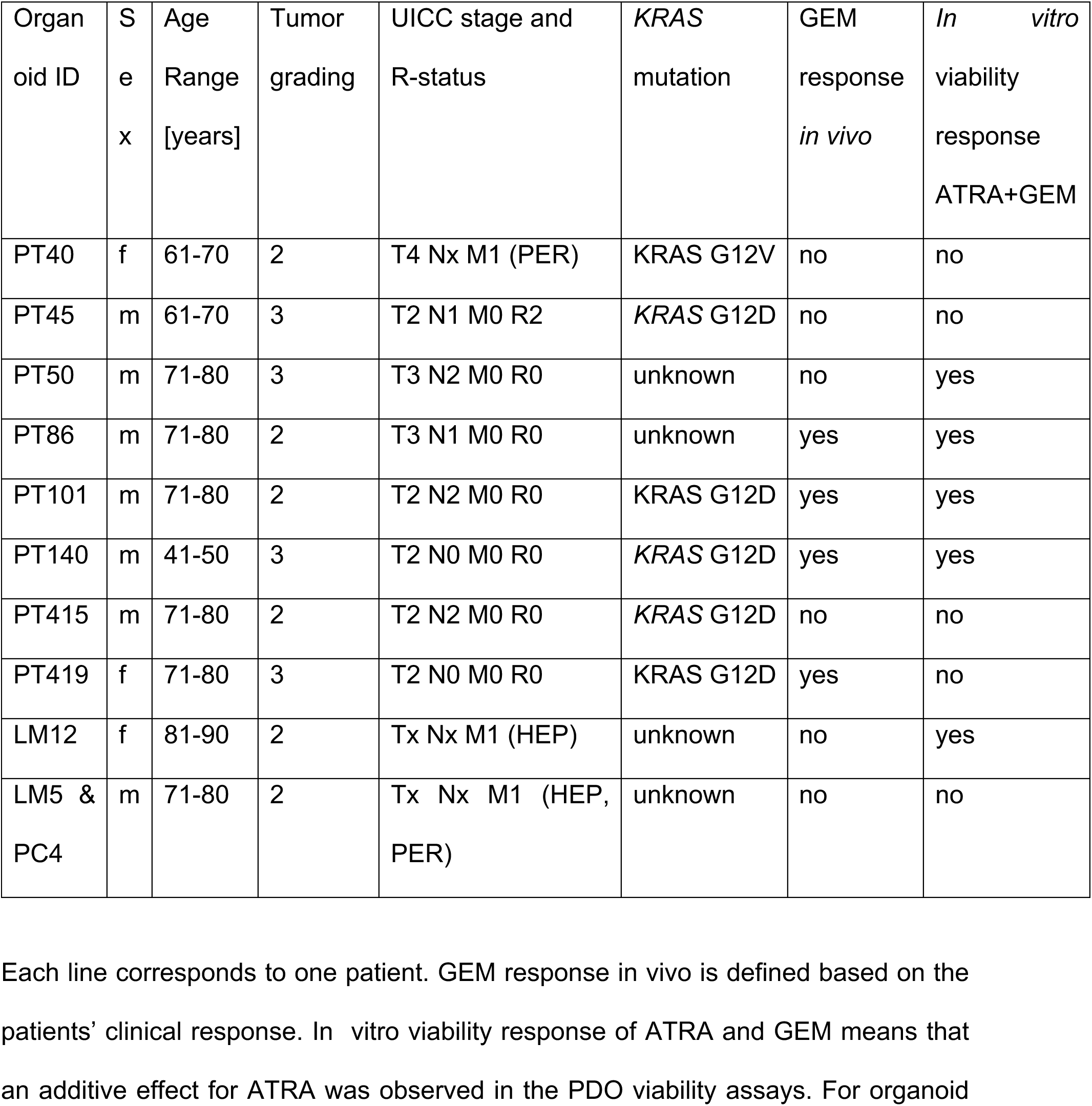
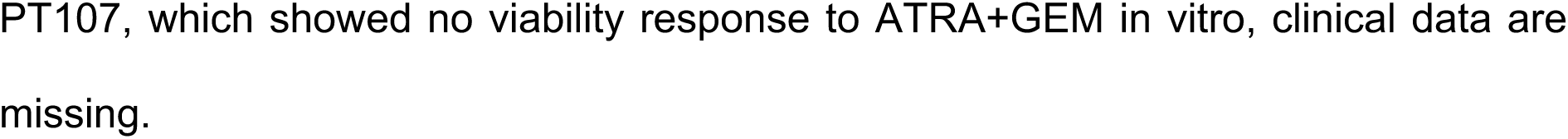
Clinicopathological data of patients who provided tissue for patient-derived organoid (PDO) cultures used in this study.

| Organoid ID | Sex | Age Range [years] | Tumor grading | UICC stage and R-status | KRAS mutation | GEM response *in vivo* | *In vitro* viability response ATRA+GEM |
| --- | --- | --- | --- | --- | --- | --- | --- |
| PT40 | f | 61-70 | 2 | T4 Nx M1 (PER) | KRAS G12V | no | no |
| PT45 | m | 61-70 | 3 | T2 N1 M0 R2 | KRAS G12D | no | no |
| PT50 | m | 71-80 | 3 | T3 N2 M0 R0 | unknown | no | yes |
| PT86 | m | 71-80 | 2 | T3 N1 M0 R0 | unknown | yes | yes |
| PT101 | m | 71-80 | 2 | T2 N2 M0 R0 | KRAS G12D | yes | yes |
| PT140 | m | 41-50 | 3 | T2 N0 M0 R0 | KRAS G12D | yes | yes |
| PT415 | m | 71-80 | 2 | T2 N2 M0 R0 | KRAS G12D | no | no |
| PT419 | f | 71-80 | 3 | T2 N0 M0 R0 | KRAS G12D | yes | no |
| LM12 | f | 81-90 | 2 | Tx Nx M1 (HEP) | unknown | no | yes |
| LM5 & PC4 | m | 71-80 | 2 | Tx Nx M1 (HEP, PER) | unknown | no | no |

Each line corresponds to one patient. GEM response *in vivo* is defined based on the patients' clinical response. *In vitro* viability response of ATRA and GEM means that an additive effect for ATRA was observed in the PDO viability assays. For organoid
PT107, which showed no viability response to ATRA+GEM in vitro, clinical data are missing.

**Supplementary Table 2:**
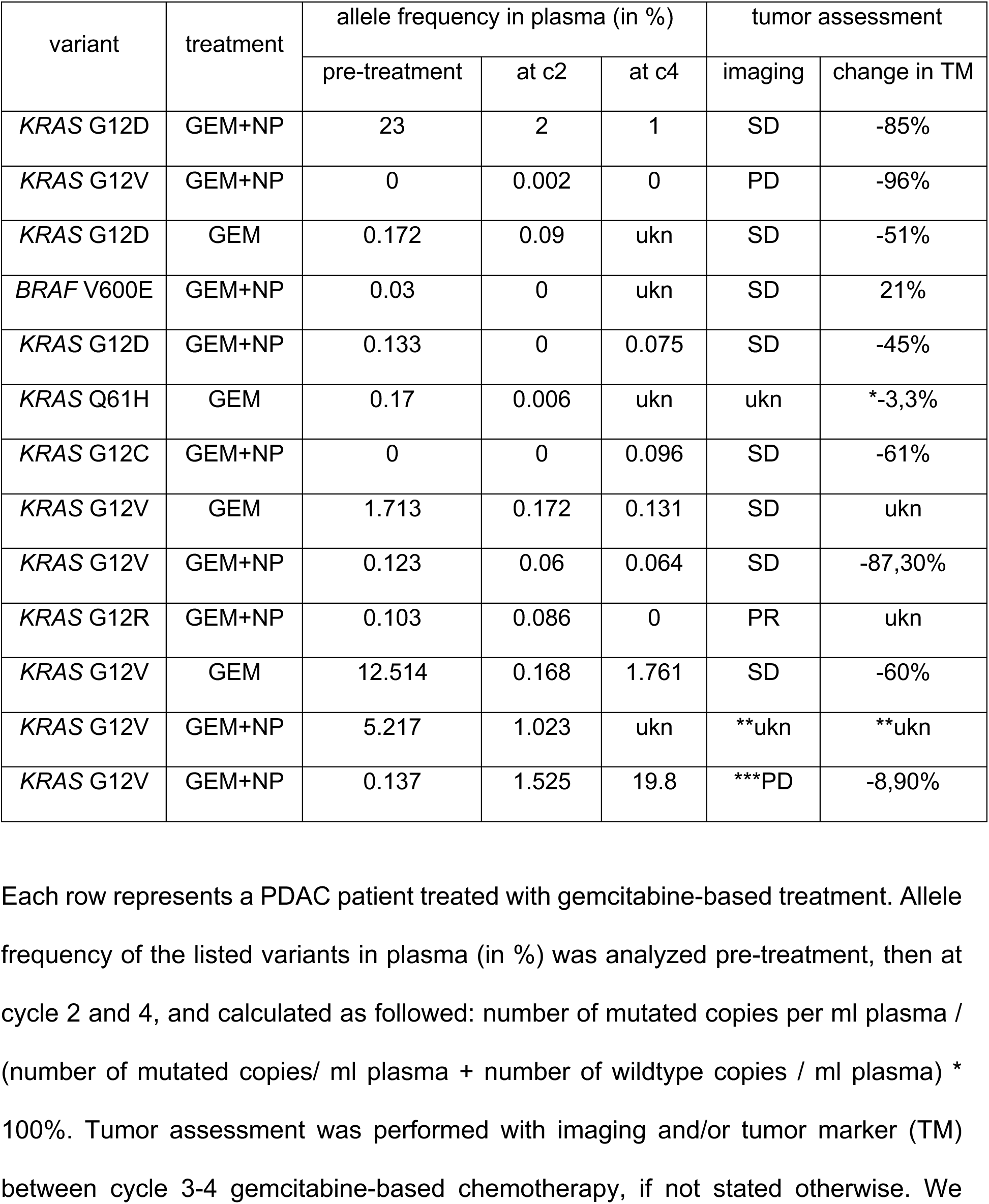
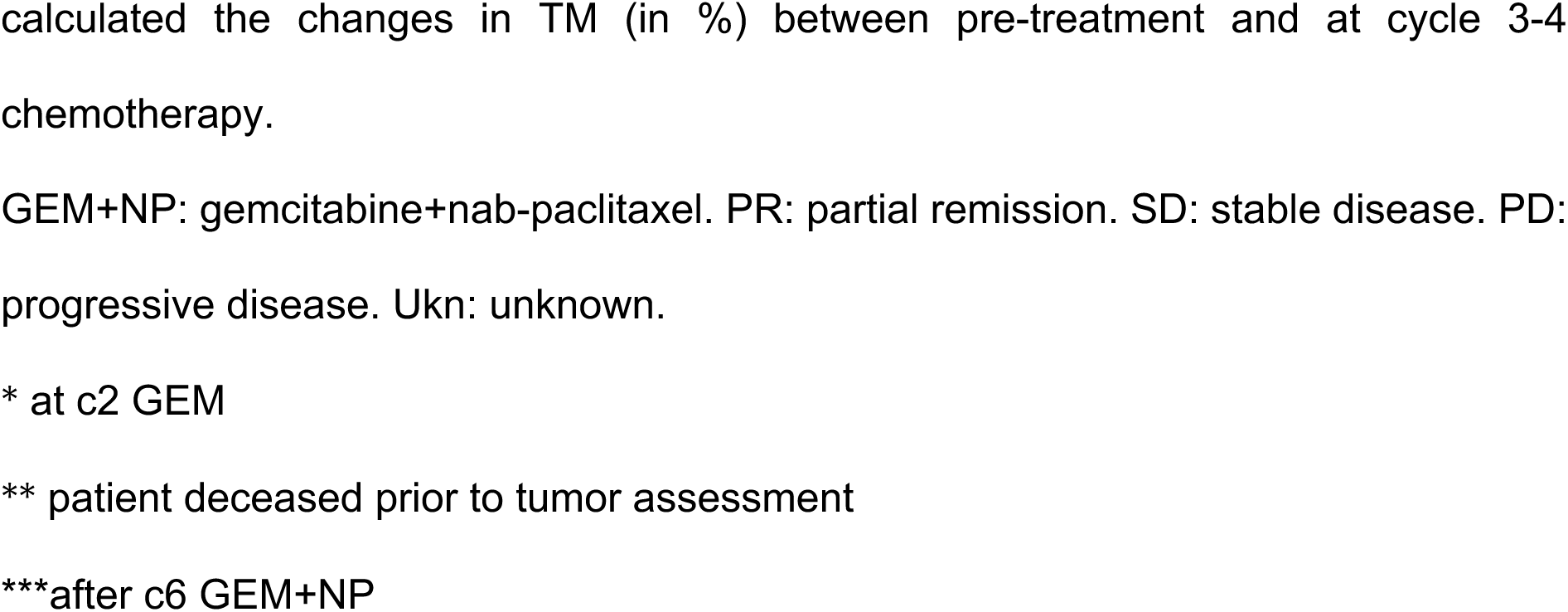
ctDNA before start of gemcitabine-based chemotherapy, at cycle 2 and 4.

| variant | treatment | allele frequency in plasma (in %) |  |  | tumor assessment |  |
| --- | --- | --- | --- | --- | --- | --- |
|  |  | pre-treatment | at c2 | at c4 | imaging | change in TM |
| *KRAS* G12D | GEM+NP | 23 | 2 | 1 | SD | -85% |
| *KRAS* G12V | GEM+NP | 0 | 0.002 | 0 | PD | -96% |
| *KRAS* G12D | GEM | 0.172 | 0.09 | ukn | SD | -51% |
| *BRAF* V600E | GEM+NP | 0.03 | 0 | ukn | SD | 21% |
| *KRAS* G12D | GEM+NP | 0.133 | 0 | 0.075 | SD | -45% |
| *KRAS* Q61H | GEM | 0.17 | 0.006 | ukn | ukn | *-3,3% |
| *KRAS* G12C | GEM+NP | 0 | 0 | 0.096 | SD | -61% |
| *KRAS* G12V | GEM | 1.713 | 0.172 | 0.131 | SD | ukn |
| *KRAS* G12V | GEM+NP | 0.123 | 0.06 | 0.064 | SD | -87,30% |
| *KRAS* G12R | GEM+NP | 0.103 | 0.086 | 0 | PR | ukn |
| *KRAS* G12V | GEM | 12.514 | 0.168 | 1.761 | SD | -60% |
| *KRAS* G12V | GEM+NP | 5.217 | 1.023 | ukn | **ukn | **ukn |
| *KRAS* G12V | GEM+NP | 0.137 | 1.525 | 19.8 | ***PD | -8,90% |

Each row represents a PDAC patient treated with gemcitabine-based treatment. Allele frequency of the listed variants in plasma (in %) was analyzed pre-treatment, then at cycle 2 and 4, and calculated as followed: number of mutated copies per ml plasma / (number of mutated copies/ ml plasma + number of wildtype copies / ml plasma) \* 100%. Tumor assessment was performed with imaging and/or tumor marker (TM) between cycle 3-4 gemcitabine-based chemotherapy, if not stated otherwise. We
calculated the changes in TM (in %) between pre-treatment and at cycle 3-4 chemotherapy. GEM+NP: gemcitabine+nab-paclitaxel. PR: partial remission. SD: stable disease. PD: progressive disease. Ukn: unknown. \* at c2 GEM \*\* patient deceased prior to tumor assessment \*\*\*after c6 GEM+NP

---

**Supplementary Figure 1:**
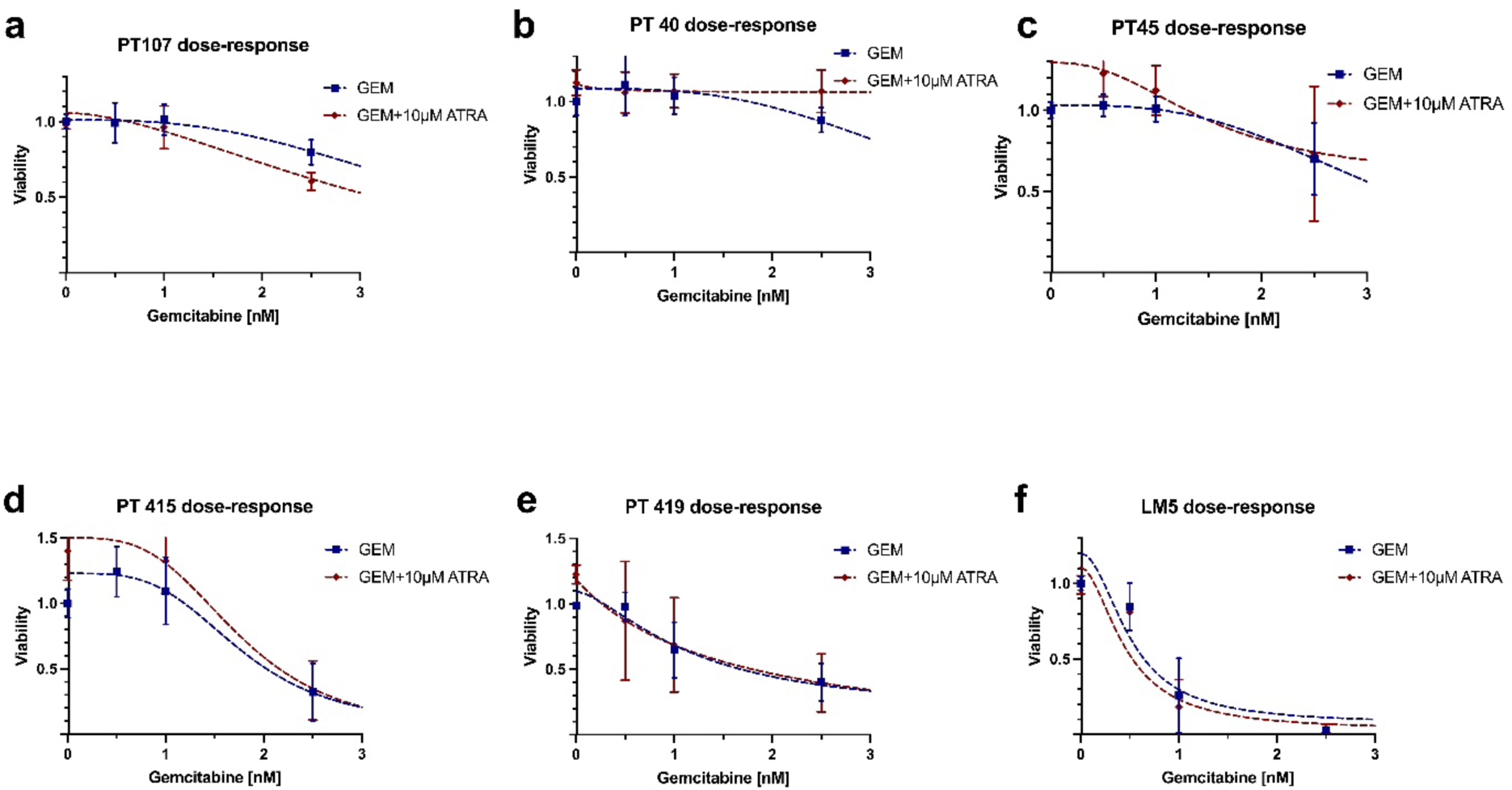
PDOs without additive effect of gemcitabine and ATRA. Dose–response curves of six PDAC PDOs treated with GEM alone or combined with 10 µM ATRA without enhanced response to ATRA combination. PDOs were exposed to increasing GEM concentrations (0.5-2.5 nM; up to 5.0 nM in selected models), as indicated. Viability was normalized to DMSO controls. Data points represent mean ± SD. Dashed lines indicate nonlinear regression fits.

**Supplementary Figure 2:**
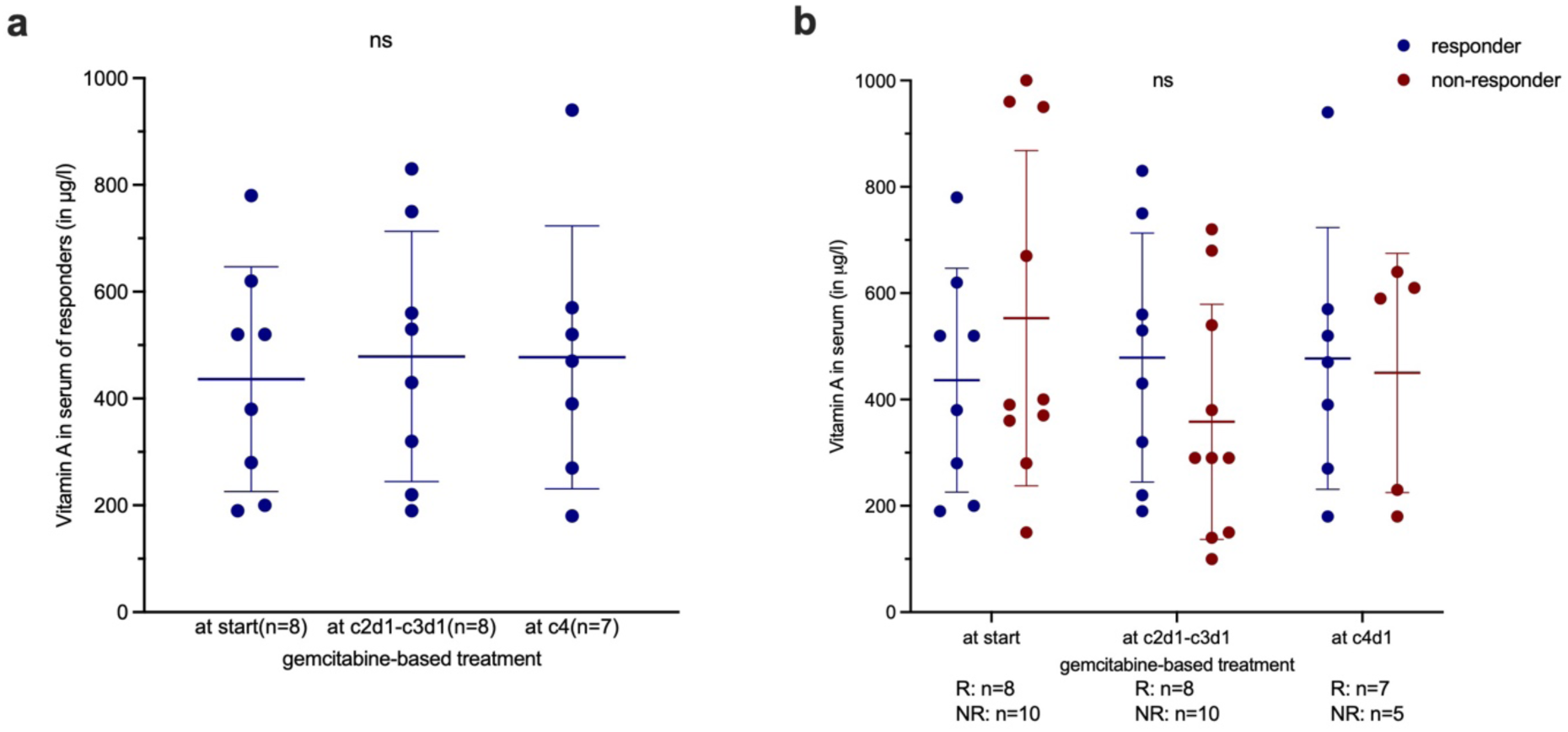
Vitamin A in serum during gemcitabine-based treatment. **a)** Serum vitamin A levels remain stable in responder during chemotherapy. Each data point represents one measurement of vitamin A in one patient. Data are expressed as mean and SD. Statistics were performed with Wilcoxon test. **b)** Serum vitamin A levels in responders (blue dots) and non-responders (red dots) during chemotherapy. Data are expressed as mean and SD. Each dot represents one patient. Comparison of responder to non-responder was performed with Mann-Whitney test.

**Supplementary Figure 3:**
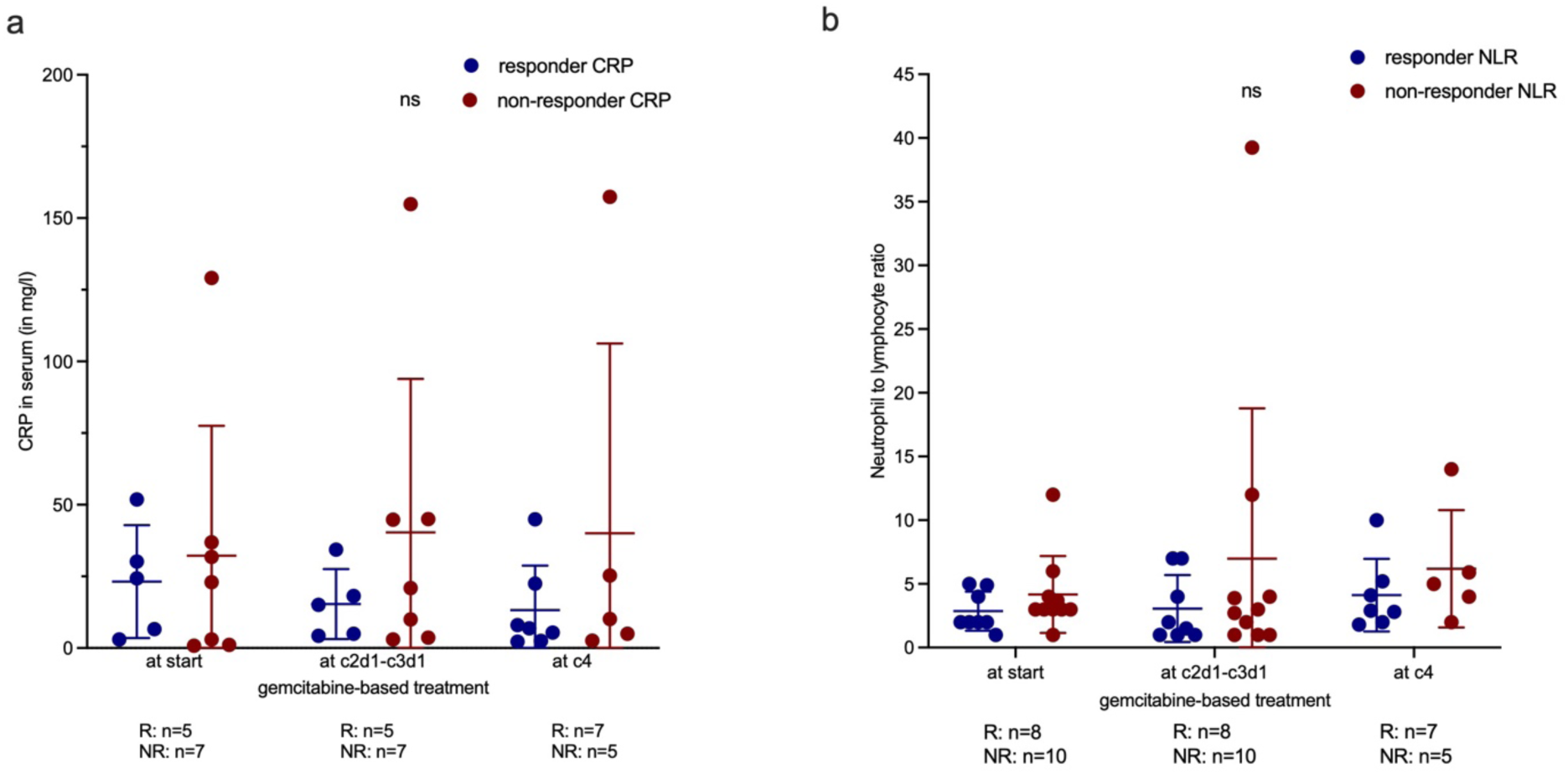
Serum CRP and NLR in responder and non-responder. **a)** Serum c-reactive protein (CRP) levels during the first four cycles GEM-based treatment in responders (blue dots) comparing to non-responders (red dots). Each dot represents one measurement in one patient. Data are shown as mean and SD. Statistic was performed with Mann-Whitney test. **b)** Neutrophil to lymphocyte ratio (NLR) in peripheral blood of responders (blue) vs. non-responders (red) to GEM-based therapy. NLR was calculated as followed: (absolute neutrophil count, x10^6^ cells/L) / (absolute lymphocyte count, x10^6^ cells/L). Data are shown as mean and SD. Statistic was performed with Mann-Whitney test.

**Supplementary Figure 4:**
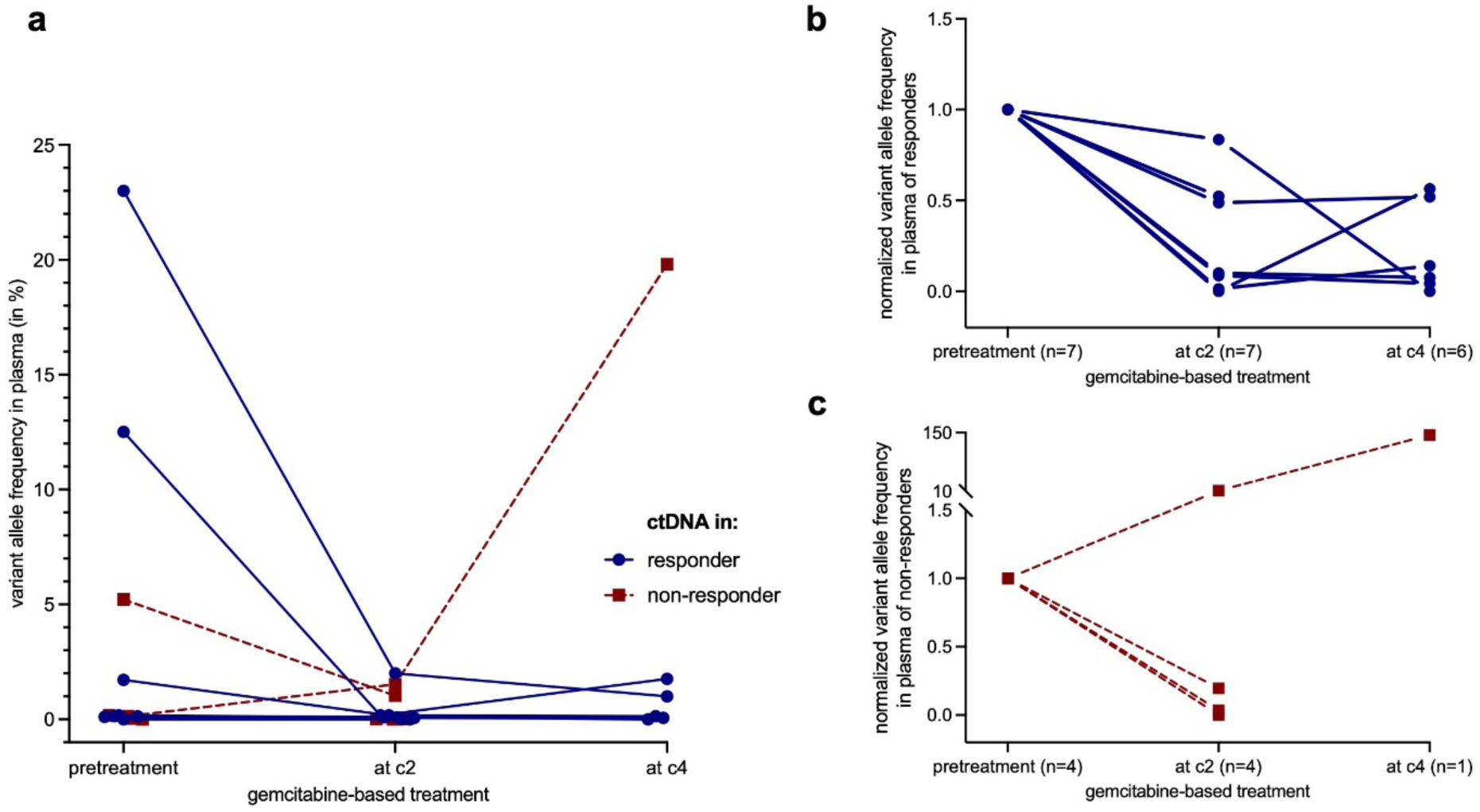
ctDNA before and during chemotherapy in PDAC patients. **a**) Variant allele frequencies (in %) of each patient before and during GEM-based chemotherapy. Blue dots are responders, red squares non-responders. *KRAS* variant was monitored in 12 patients, *BRAF V600E* in 1 patient. Each patient had one measurement. **b-c)** Plasma ctDNA levels of responders (blue, **b**) and non-responders (red, **c**) pre-treatment and during gemcitabine-based chemotherapy are shown. We monitored *KRAS* variants in plasma of 12 patients and *BRAF V600E* in 1 patient. For each patient, allele frequencies (in percent) of each timepoint were normalized to the baseline value (=1). 2 patients with no detectable ctDNA at baseline (AF=0%) were excluded.

**Supplementary Figure 5:**
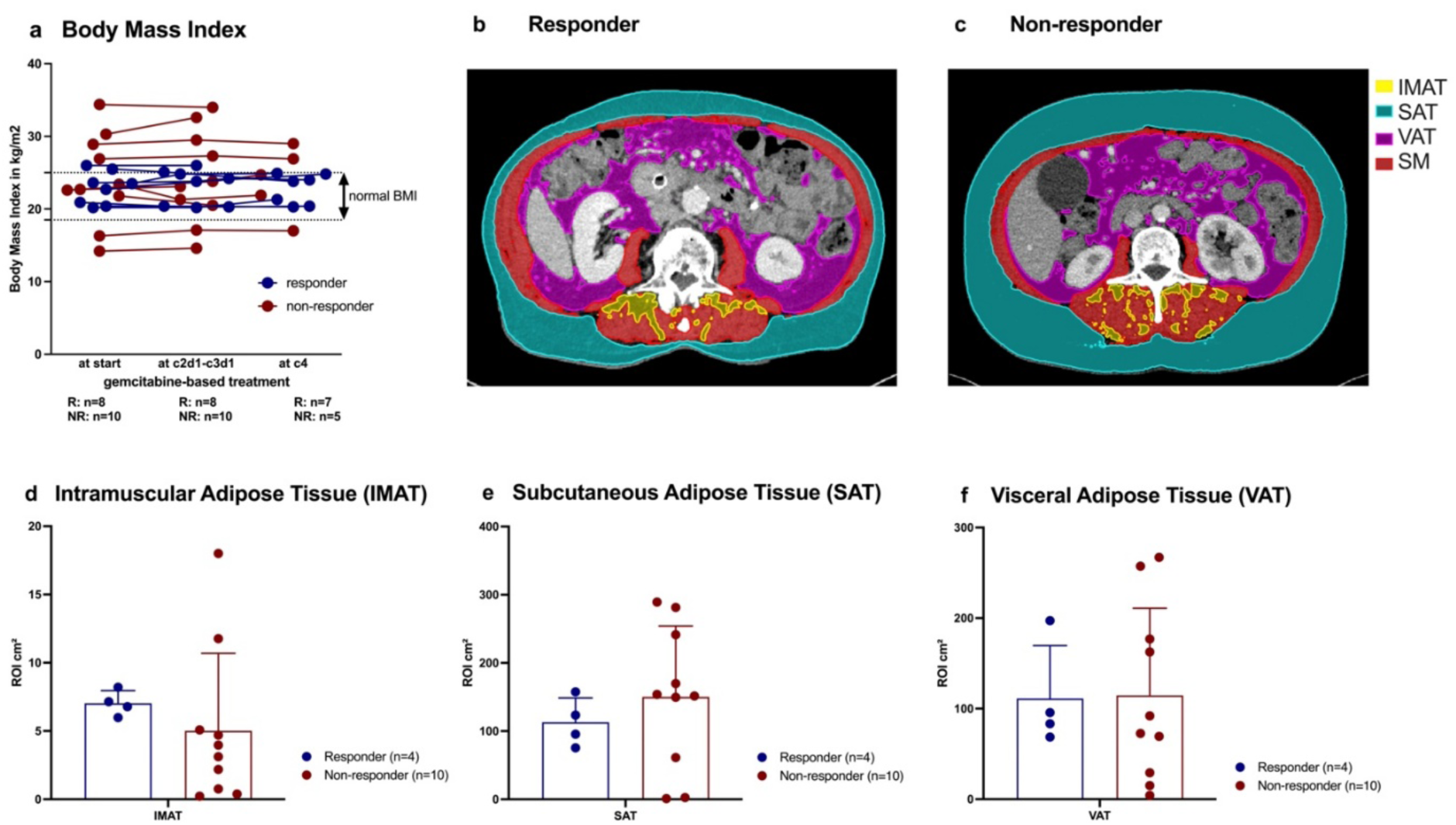
Body mass index and body composition during chemotherapy. **a)** Body Mass Index (BMI) in kg/m^2^ of responder (blue) vs. non-responder (red) to GEM-based treatment at baseline and then during chemotherapy. Values between the lines at 18.5 and 25 kg/m^2^ are in the normal BMI range. **b, c)** Example CT-scans with marked intramuscular adipose tissue (IMAT; yellow), subcutaneous adipose tissue (SAT; turquoise), visceral adipose tissue (VAT; pink) and skeletal muscle (SM; red) are shown for a responder in **b)** and a non-responder in **c)**. Baseline d) IMAT, **e)** SAT and **f)** VAT of responder (blue) vs. non-responder (red) were defined as region of interest (ROI, in cm^2^), at the 3rd lumbar vertebrae, before start of treatment using NORA software based on routine CT scans. The differences between the groups are not significant. The variance homogeneity for IMAT is significantly different between the two groups (f-test, p<0.05).

**Supplementary Figure 6:**
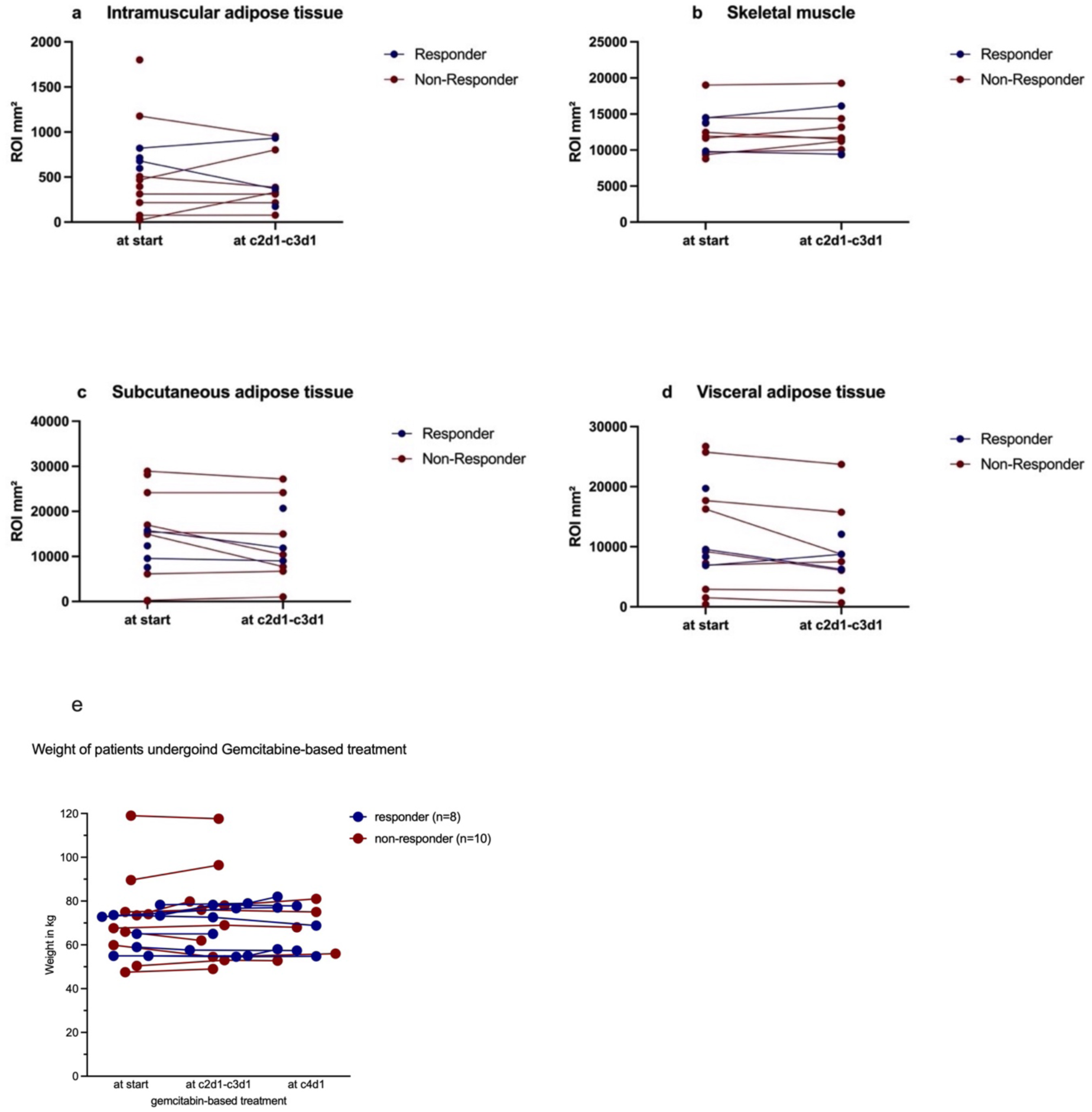
Body composition and weight over time in responder and non-responder of GEM-based treatment. **a)** Intramuscular adipose tissue (IMAT), **b)** skeletal muscle (SM), **c)** subcutaneous adipose tissue (SAT) and **d)** visceral adipose tissue (VAT) defined as region of interest (ROI, in mm^2^), at the 3rd lumbar vertebrae in routine CT-scans at baseline and then during chemotherapy. Responder to GEM-based treatment are marked blue and non-responder red. The defined values remain stable during chemotherapy. **e)** Weight of patients undergoing gemcitabine-based treatment at baseline, cycle 2 and 4.

**Supplementary Figure 7:**
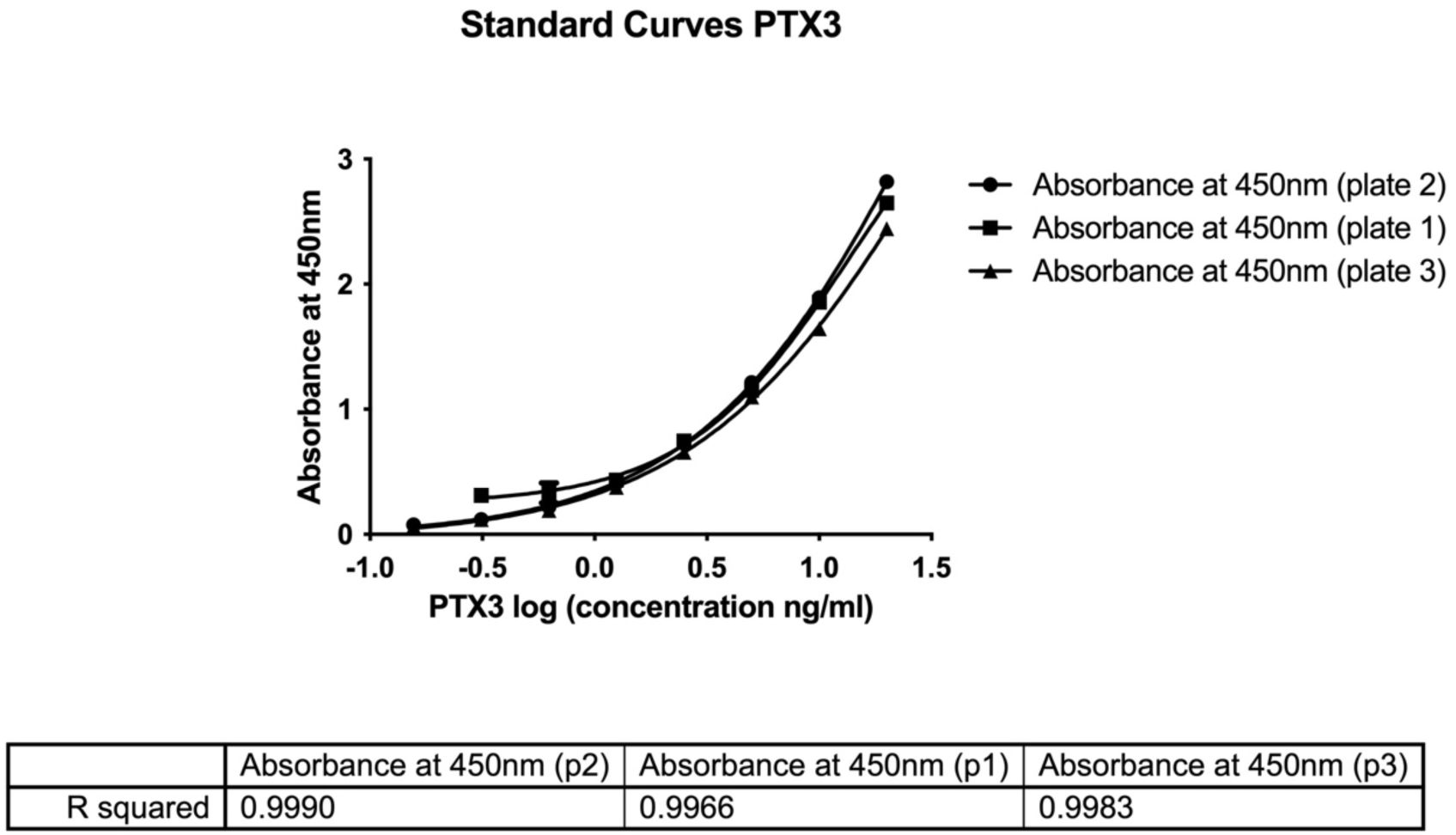
Standard Curves for the PTX3 ELISA Assay. Polynomial regression graphs were constructed for standard curves for PTX3 analysis with ELISA. Plates were read at 450nm.

